# Reactivation of herpesvirus type-6 and IgA/IgM-mediated responses to activin-A underpin Long COVID, including affective symptoms and chronic fatigue syndrome

**DOI:** 10.1101/2023.07.23.23293046

**Authors:** Aristo Vojdani, Abbas F. Almulla, Bo Zhou, Hussein K. Al-Hakeim, Michael Maes

## Abstract

**Abstract:** *Background:* Persistent infection with severe acute respiratory syndrome coronavirus 2 (SARS-CoV-2), reactivation of dormant viruses, and immune-oxidative responses are involved in Long COVID.

*Objectives:* To investigate whether Long COVID and depressive, anxiety and chronic fatigue syndrome (CFS) symptoms, are associated with IgA/IgM/IgG to SARS-CoV-2, human Herpesvirus type 6 (HHV-6), Epstein-Barr Virus (EBV), and immune-oxidative biomarkers.

*Methods:* We examined 90 Long COVID patients and 90 healthy controls. We measured serum IgA/IgM/IgG against HHV-6 and EBV and their deoxyuridine 5′-triphosphate nucleotidohydrolase (duTPase), SARS-CoV-2, and activin-A, C-reactive protein (CRP), advanced oxidation protein products (AOPP), and insulin resistance (HOMA2-IR).

*Results:* Long COVID patients showed significant elevations in IgG/IgM-SARS-CoV-2, IgG/IgM-HHV-6 and HHV-6-duTPase, IgA/IgM-activin-A, CRP, AOPP, and HOMA2-IR. Neural network analysis yielded a highly significant predictive accuracy of 80.6% for the Long COVID diagnosis (sensitivity: 78.9%, specificity: 81.8%, area under the ROC curve=0.876); the topmost predictors were: IGA-activin-A, IgG-HHV-6, IgM-HHV-6-duTPase, IgG-SARS-CoV-2, and IgM-HHV-6 (all positively) and a factor extracted from all IgA levels to all viral antigens (inversely). The top-5 predictors of affective symptoms due to Long COVID were: IgM-HHV-6-duTPase, IgG-HHV-6, CRP, education, IgA-activin-A (predictive accuracy of r=0.636). The top-5 predictors of CFS due to Long COVID were in descending order: CRP, IgG-HHV-6-duTPase, IgM-activin-A, IgM-SARS-CoV-2, and IgA-activin-A (predictive accuracy: r=0.709).

*Conclusion:* Reactivation of HHV-6, SARS-CoV-2 persistence, and autoimmune reactions to activin-A combined with activated immune-oxidative pathways play a major role in the pathophysiology of Long COVID as well as the severity of affective symptoms and CFS due to Long COVID.

## Introduction

Between three and six months following acute COVID-19 infection, many individuals may acquire long coronavirus (Long COVID) or post-COVID illness (Groff, Sun et al. 2021). Despite the wide range of symptomatology, chronic fatigue, symptoms of depression and anxiety, problems with neurocognition, shortness of breath, and gastrointestinal dysfunctions are the most common Long COVID symptoms (Renaud-Charest, Lui et al. 2021, Sandler, Wyller et al. 2021, Titze-de-Almeida, da Cunha et al. 2022). We found that fatigue, physiosomatic, depressive, and anxiety symptoms are manifestations of a single core (latent vector) referred to as the physio-affective phenome of acute COVID-19 infection and Long COVID (Al-Hadrawi, Al-Rubaye et al. 2022, Al-Jassas, Al-Hakeim et al. 2022, Almulla, Al-Hakeim et al. 2023). Based on the results of the Hamilton Depression (HAMD) and Anxiety (HAMA) Rating Scales, the Fibromyalgia and Fatigue (FF) Rating Scale scores, this physio-affective phenome was created using a precision nomothetic technique (Maes 2022, Maes and Stoyanov 2022).

We discovered that abnormalities in the immune-inflammatory (as indicated by higher C-reactive protein or CRP, and NLRP3 inflammasome biomarkers), oxidative stress (as indicated by increased advanced oxidation protein products (AOPP), as well as increased insulin resistance (IR) partially explain the physio-affective phenome of acute and Long COVID (Al-Jassas, Al-Hakeim et al. 2022, Almulla, Al-Hakeim et al. 2023). We demonstrated in our Long COVID studies that decreased oxygen saturation (SpO2) and elevated peak body temperature (PBT) during the acute infection stage are related to persistent fatigue and depressive symptoms many months later (Al-Hadrawi, Al-Rubaye et al. 2022). Increased PBT and decreased SpO2, both of which are linked to higher morbidity, are indicators of the severity of the inflammatory response during the acute phase of disease (Al-Jassas, Al-Hakeim et al. 2022). Therefore, the severity of the acute infectious phase contributes to aberrations in immune and oxidative pathways, with low SpO2 linked to changes in oxidative stress pathways, and increased PBT linked to increased CRP and decreased antioxidant defenses (Al-Hakeim, Al-Rubaye et al. 2022).

Nevertheless, the precise pathophysiology of Long COVID and the mechanisms underlying the activated immune pathways in Long COVID, have not yet been clarified. Regular detection of SARS-CoV-2 viral RNA or antigens in blood, feces, and other tissue samples up to many months after the initial diagnosis of COVID-19 led to the assumption that persistent SARS-CoV-2 infection may be involved (Li, Li et al. 2021, Cheung, Goh et al. 2022, Natarajan, Zlitni et al. 2022, Rahmani, Dini et al. 2022). According to (Chen, Julg et al. 2023, Vojdani, Vojdani et al. 2023, Yang, Zhao et al. 2023), this viral persistence may cause the immune system to remain persistently active, which would then contribute to the ongoing neuropsychiatric symptoms and chronic fatigue syndrome (CFS) reported in people with Long COVID.

A growing body of research also demonstrates that acute and critical SARS-CoV-2 infection can trigger the reactivation of other viruses, such as the Epstein-Barr Virus (EBV), Human Herpesvirus type 6 (HHV-6), and other viruses (Balc’h, Pinceaux et al. 2020, Lehner, Klein et al. 2020). A high proportion of patients with Long COVID show IgG titers that are positive for the EBV reactivation markers early antigen-diffuse (EA-D) and small viral capsid antigen p23 (Gold, Okyay et al. 2021, Jon, Jamie et al. 2022). It is not yet known, however, if these viruses contribute to the development of neuropsychiatric disorders and signs of chronic fatigue syndrome (CFS) during Long COVID (Proal and VanElzakker 2021).

There may be several mechanisms explaining why reactivation of these viruses increases susceptibility to neuropsychiatric symptoms and CFS. First, the viruses listed above might make it easier for SARS-CoV-2 to enter cells (Vojdani, Vojdani et al. 2023). Second, according to several studies (Gold, Okyay et al. 2021, Zubchenko, Kril et al. 2022, Vojdani, Vojdani et al. 2023), these viruses probably lead to mitochondrial breakdown and dramatically change energy metabolism. Since mitochondrial dysfunctions have been linked to the pathobiology of immune-mediated illnesses such as CFS and affective disorders, a same mechanism could lead to Long COVID (Morris and Maes 2014, Anderson, Almulla et al. 2023). Third, while a small number of studies have investigated the presence of immunoglobulins (IgA, IgM, and IgG) against persistent SARS-CoV-2 and reactivated viruses, no studies have investigated its deoxyuridine 5′-triphosphate nucleotidohydrolase (duTPase), a nucleotide metabolic enzyme that is an important regulator of the innate and adaptive immune responses (Vojdani, Vojdani et al. 2023). The release of the latter in exosomes may contribute to CFS by increasing the synthesis of cytokines, activin-A, plasma cells, and antibodies, and consequently abnormal immunological responses (Cox, Alharshawi et al. 2022). The immune responses against human growth proteins like activin-A, interferon (IFN)-α2, a master cytokine in type 1 IFN signaling, heat shock protein (HSP)60, and HSP90, which support proteostasis and regulate apoptosis and the cell cycle, are not studied in relation to neuropsychiatric symptoms and CFS caused by Long COVID (Hu, Yang et al. 2022). The etiology of depressive symptoms and CFS is associated with changes in activin-A, HSP60 and HSP90 (Dow, Russell et al. 2005, Ageta, Murayama et al. 2008, Zheng, Link et al. 2017, Sagulkoo, Plaimas et al. 2022). However, there is little or no information on the relationship between altered IgA/IgG/IgM responses to activin-A, IFN-α2, HSP-60, and HSP90 and affective symptoms and CFS caused by Long COVID.

Hence, the aim of the current study is to compare the levels of IgA/IgG/IgM antibodies against SARS-CoV-2, HHV-6, and HHV-6-dutPase, EBV and EBV-duTPase, activin-A, HSP60, HSP90, and IFN-α2 in Long COVID patients to those of healthy persons. We also want to see if the severity of the affective symptoms and CFS due to Long COVID is predicted by these IgA/IgG/IgM levels.

## Participants and Methods

### Participants

The participants in this study included 90 Long COVID patients and 90 healthy controls. Senior clinicians diagnosed the patients, using criteria specified by the World Health Organization (WHO, 2021). According to these guidelines, a Long COVID patient must satisfy the following criteria: a confirmed COVID-19 infection; at least two symptoms, such as fatigue, memory or concentration issues, muscle aches, loss of smell or taste, emotional distress, and cognitive impairment that interfere with daily life; symptoms persisting after the acute phase of the illness or appearing 2-3 months later; and the continuity of these symptoms for at least two months, even 3-4 months after the acute phase of the illness. The research strategy employed two study designs. The first part (n=72) was a case-control and retrospective cohort study to determine the influence of PBT, SpO2, and duration of the acute infection on the biomarkers and the effects of the biomarkers on the symptoms of Long COVID. The second study (n=180) was a case control study of the associations between biomarkers and Long COVID versus healthy controls. Group 1 consisted of Iraqi participants. Acute COVID-19 infection was diagnosed by virologists and specialist physicians. When determining the prognosis of Long COVID, the following factors were taken into account: a) presence of symptoms of Long COVID (see above) that lasted for 12-16 weeks or longer; b) the acute infectious phase is characterized by severe infection symptoms, including fever, cough, shortness of breath, and the loss of senses of smell and taste; c) during the acute phase of illness, a positive test result for reverse transcription real-time polymerase chain reaction (rRT-PCR) and immunoglobulin M (IgM) antibodies against SARS-CoV-2. Patients in subgroup 1 were treated at a variety of hospitals, including the Imam Sajjad Hospital, the Hassan Halos Al-Hatmy Hospital for Transmitted Diseases, the Middle Euphrates Center for Cancer, the Al-Najaf Teaching Hospital, and the Al-Sader Medical City of Najaf, Iraq. In Najaf, controls were recruited from the same catchment area. Controls were excluded if they exhibited symptoms suggestive of COVID-19 or another infectious disease, a positive rRT-PCR test, or elevated IgM antibodies against SARS-CoV-2. Participants were excluded if they had a lifetime history of a major depressive episode, bipolar disorder, dysthymia, generalized anxiety disorder, panic disorder, schizo-affective disorder, schizophrenia, psycho-organic syndrome, or substance use disorder (SUD) (except for nicotine dependence). This investigation excluded patients with neuro immune, autoimmune, and immune disorders including Parkinson’s disease, CFS, Alzheimer’s disease, multiple sclerosis, stroke, psoriasis, chronic kidney disease, COPD, and scleroderma. Women who were pregnant or nursing were excluded. Subgroup 2 consisted of 32 Long COVID patients who were examined at the Cyrex Laboratory in California, United States. These patients exhibited symptoms of fatigue, brain fog, neurocognitive decline, shortness of breath, headaches or vertigo, difficulty sleeping, cough, chest pain, joint or muscle pain, gastro-intestinal disorders, and irregular menstruation that persisted for at least 12 weeks following the initial infection with COVID-19. All individuals were screened for elevated IgG antibodies against the spike protein and nucleoprotein of SARS-CoV-2. In addition, 76 pre-COVID control sera were acquired from Innovative Research (Novi, MI). The sera were extracted from healthy volunteers and tested for the presence of HIV and hepatitis C. Using the SARS-CoV-2 IgG antibody assay by Zeus Scientific, all 76 sera were tested for the presence of SARS-CoV-2 antibody and were found to be negative. All sera were stored at -20°C until their use in the antibody assays.

The College of Medical Technology Ethics Committee of the Islamic University of Najaf in Iraq (Document No. 34/2023) granted ethical approval for this investigation. Before participating in our study, all participants or their legal guardians gave their written consent. The International Conference on Harmonization of Good Clinical Practice, the Belmont Report, the Council of International Organizations of Medicine (CIOMS) Guideline, as well as Iraqi and international ethical and privacy statutes, were all adhered to in the planning and execution of this study. Our institutional review board follows the International Guidelines for the Conduct of Safe Human Research (ICH-GCP).

### Clinical measurements

Approximately three to four months after recovering from acute COVID-19, all participants in study 1 were interviewed by an experienced psychiatrist. The purpose of this interview was to collect information regarding the sociodemographic and clinical characteristics of the participants. The psychiatrist evaluated the presence and severity of CFS symptoms using the Fibro-fatigue (FF) scale (Zachrisson, Regland et al. 2002). The Hamilton Depression Rating Scale (HAMD) (Hamilton 1960) and the Beck Depression Inventory-II (BDI-II) (Hautzinger 2009) were used to measure the severity of depression. To evaluate the severity of anxiety symptoms, the Hamilton Anxiety Rating Scale (HAMA) (Hamilton 1959) was utilized. The severity of the Long COVID phenome was estimated by computing a z unit-based composite score calculated as z HAMD + z HAMA + z BDI (labeled as comp_AFFECT). The diagnostic criteria for tobacco use disorder (TUD) were derived from the Diagnostic and Statistical Manual of Mental Disorders, Fifth Edition (DSM-5). The body mass index (BMI) was calculated by dividing the individual’s weight in kilograms by the square of their height in meters. We extracted the SpO2 and PBT values from the patients’ hospitalization records during the acute infection phase. A highly competent paramedical professional took these measurements using an electronic oximeter manufactured by Shenzhen Jumper Medical Equipment Co. Ltd. and a digital sublingual thermometer with an audible alert, respectively. The duration of the patient’s illness was deduced from their medical records.

### Assays

Blood samples were collected from fasting participants early in the morning, between 7:30 and 9:00 a.m. Venous blood (five milliliters) was drawn and placed into sterilized, clear serum tubes. Samples that showed signs of hemolysis were not used. After allowing the blood to clot for ten minutes, the samples were centrifuged for five minutes at a speed of 3000 rpm. The resulting serum was then carefully transferred into several new Eppendorf tubes. Measurements for CRP in human serum were performed using the CRP latex slide test, a product of Spinreact® (Barcelona, Spain). Serum AOPP levels were determined using ELISA kits sourced from Nanjing Pars Biochem Co., Ltd. (Nanjing, China). The Homeostatic Model Assessment for Insulin Resistance (HOMA2-IR), a measure of insulin resistance, was computed from fasting insulin and serum glucose levels using a HOMA2 calculator available at https://www.dtu.ox.ac.uk/homacalculator/.

### Antigens and Sera

SARS-CoV-2 spike protein superantigen (amino acid sequence of 2 671-700 (NH2) CASYQTQTNSPRRRARSVASQSIIAYTMSLGA (COOH)) which has some sequence and structure similarity to Staphylococcus enterotoxin B (SEB) (Cheng, Zhang et al. 2020); Epstein-Barr virus (EBV) lytic protein that is produced during reactivation of the virus (Williams, Cox et al. 2016), amino acid sequence [NH2]WAT[C(Cam)]AFEEVPGLA[M(OX)]GDSGLSEALEGR[COOH] of duTPase; human herpesvirus 6 A&B (HHV-6A&B); HHV-6 duTPase that function as pathogen associated molecular pattern proteins for TLR2 (Cox, Alharshawi et al. 2022) amino acid sequence of (NH2) CHGLLIETYIWNKDTIPSIKIFNST (COOH); human molecular chaperones, HSP60 (amino acid sequence (NH2) AEIPKEEVKPFITESKPSVEQRKQDDKK (COOH)) and HSP90 (amino acid sequence (NH2) KTFPPTEPKKDKKKKADETQALPQRQKKQQ(COOH)) which share antigenic epitopes with SARS-CoV-2, were synthesized with purity of greater than 85% by Bio Synthesis (Lewisville, TX, USA). Citrullinated EBV EBNA2 peptide (sequence CQGR<u>C</u>GRWRG-cit-GRSKGRG<u>C</u>RMH) was synthesized by Bio-Synthesis (Lewisville, TX, USA) (Trier, Holm et al. 2018). Recombinant human interferon alpha-2 (IFN-α2) protein was purchased from Novus Biologicals LLC (CO, USA). Human activin-A was purchased from ProSpec (Rehovot, Israel). Recombinant human activin-A Ab151687 was purchased from Abcam (Cambridge, MA, USA).

### Antibody measurements

We used an enzyme-linked immunosorbent assay (ELISA) to measure the presence of antibodies (IgA, IgG, IgM) against the SARS superantigen, HHV-6, HHV-6-duTPase, Citrullinated EBV EBNA2 peptide, EBV-duTPase, HSP60 and HSP90, IFN-α2, and activin-A. SARS superantigen and HHV-6 were synthesized and provided with purity greater than 85% by Bio-Synthesis (Lewisville, TX, USA). Additionally, we obtained activin-A from ProSpec (Rehovot, Israel). The procedure was as follows: a) A Tris buffer with a pH of 7.2 was used to dissolve the proteins and peptides. Then, in a 0.1 M carbonate buffer at pH 9.6, we put 100 ml of each at concentrations ranging from 0.5 to 1 microgram to a series of microwell plates, b) After 16 hours in room temperature (25°C) incubation, the plates were subjected to refrigeration for another 8 hours, c) After removing the contents, the plates were washed three times with 250 ml of a 0.01 M phosphate buffer saline solution with a pH of 7.4, containing 0.05% Tween 20, d) We added 250 ml of a solution containing 2% bovine serum albumin (BSA) and 2% dry milk in the same PBS buffer to each well, and then incubated the plates again to prevent non specific binding of serum immunoglobulins e) After four washes in PBS buffer, we applied serum dilutions of 1:100 for IgG and IgM and 1:50 for IgA antibodies to duplicate wells of the microtiter plates, f) After incubating for 1 hour at 25°C, we washed the ELISA plates five times with the PBS buffer. We then added 100 ml of alkaline phosphatase-labeled anti-human IgG (dilution at 1:800), anti-human IgM (dilution at 1:600), and anti-human IgA (dilution at 1:200) antibodies to the appropriate sets of plates, g) After additional washing, we added 100 ml of substrate and stopped the color development by adding 50 ml of 1 N NaOH. The intensity of the color was measured using an ELISA reader at a wavelength of 405 nm. We also used several wells coated with BSA, human serum albumin (HAS), and fetal bovine serum as negative controls or blanks, and used sera from patients with ME/CSF and long COVID with known antibody titers as calibrators and positive controls. The ELISA optical densities (ODs) were converted to indices by dividing each sample OD by the calibrator OD, after subtracting the background OD from both, using the flowing formula:

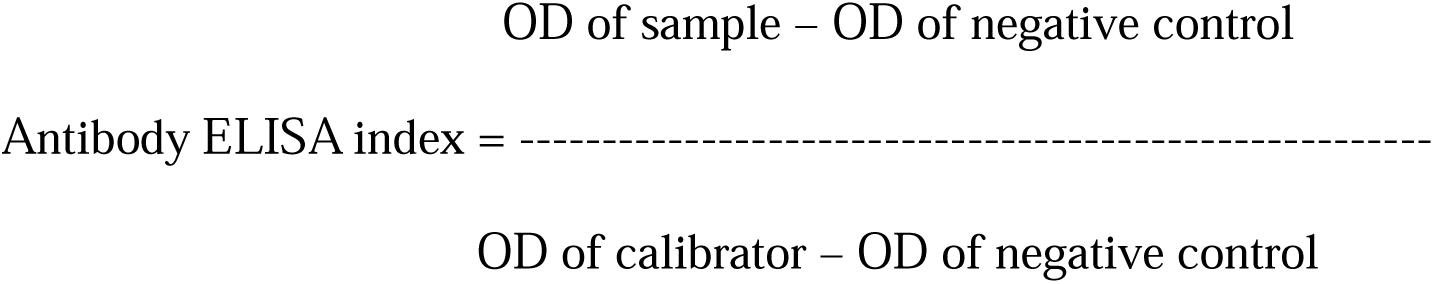

### Statistical analysis

All statistical analyses were performed using the 29th version of IBM’s SPSS Windows software. We employed analysis of variance (ANOVA) to assess differences in continuous variables across study groups. Additionally, we used contingency table analysis to investigate the relationships between categorical variables. In the total study group (n=180) we performed binary logistic regression analysis with Long COVID as dependent variable (and healthy controls as reference group) using the IgA/IgG/IgM responses as explanatory variables while adjusting for the effects of age, sex, and research site. We computed B (SE), and Wald statistics with exact p values, Odds ratio with 95 confidence intervals (CI), the classification table, and Nagelkerke pseudo-R square (used to estimate the effect size). A neural network analysis was conducted to predict the differentiation between individuals with Long COVID and control subjects, as well as the clinical continuous data. In our multilayer perceptron neural network models, we used the most significant IgG, IgM, IgA responses to the antigens, age, gender, level of education, with or without AOPP, HOMA2-IR, and CRP. An automatic feedforward network model with one or two hidden layers was constructed using up to eight nodes. The models were trained using the batch type, with 250 epochs. The stopping criterion was set to a single successive step that did not further decrease the error term. We calculated the error and the relative error, the percentage of misclassifications, and the area under the receiver operating characteristic curve (AUC ROC). The predictive accuracy of the models was estimated using the confusion matrix (for binary outputs), or the predicted versus observed coefficient of determination (R^2^). The input factors’ significance and relative importance were also assessed and displayed in an importance chart. We initially divided the participants into three categories: (a) a training group, which constituted 46.7 % of all participants and was used to determine the model’s parameters; (b) a testing group, which made up 20% of all participants and was used to avoid overfitting; and (c) a holdout group, which accounted for 33.3% of all participants and was used to assess the model’s predictive validity. We used an oversampling correction (using true observations) to compensate for a disbalanced data set when performing binary regressions and neural networks with a binary output variable. Principal component analysis (PCA) was employed for feature reduction purposes. The PC was accepted as a validated concept when the explained variance (EV) was at least 50%, the anti-image correlation matrix was adequate, the factoriability metrics were adequate with the Kaiser-Meyer-Olkin (KMO) metric > 0.7, and Bartlett’s sphericity test were significant, and PC loadings > 0.7. The significance level for all tests was set at a p-value of 0.05, and all tests were two-tailed. A priori power analysis (G*Power 3.1.9.7.) shows that the estimated sample size should be at least 126 to detect differences in a chi square test with an effect size of 0.25 given p=0.05, power=0.8 and df=1.

## Results

### Sociodemographic data, psychological measurements, and immunological parameters

The current study presents sociodemographic data, PBT, SpO2 and duration of the acute phase in **Table 1**. This table also encompasses scores from several psychological and biomarker assessments, including the FF, HAMA, HAMD, BDI, CRP, AOPP and HOMA2-IR levels. Sociodemographic data (except residency) such as age, gender, BMI, marital state, smoking, and education did not significantly differ among study groups. We found that PBT was significantly increased and SpO2 significantly decreased during the acute infectious phase in Long COVID patients versus controls. Our results also revealed that Long COVID patients show significantly elevated scores of FF, HAMA, BDI and HAMD. Additionally, CRP, AOPP, and HOMA2IR levels were significantly higher in Long COVID patients as compared to controls. In the Long COVID patients, the mean (±SD) duration of acute infectious illness was 14.1 (±5.6) days.

**Table 1:**
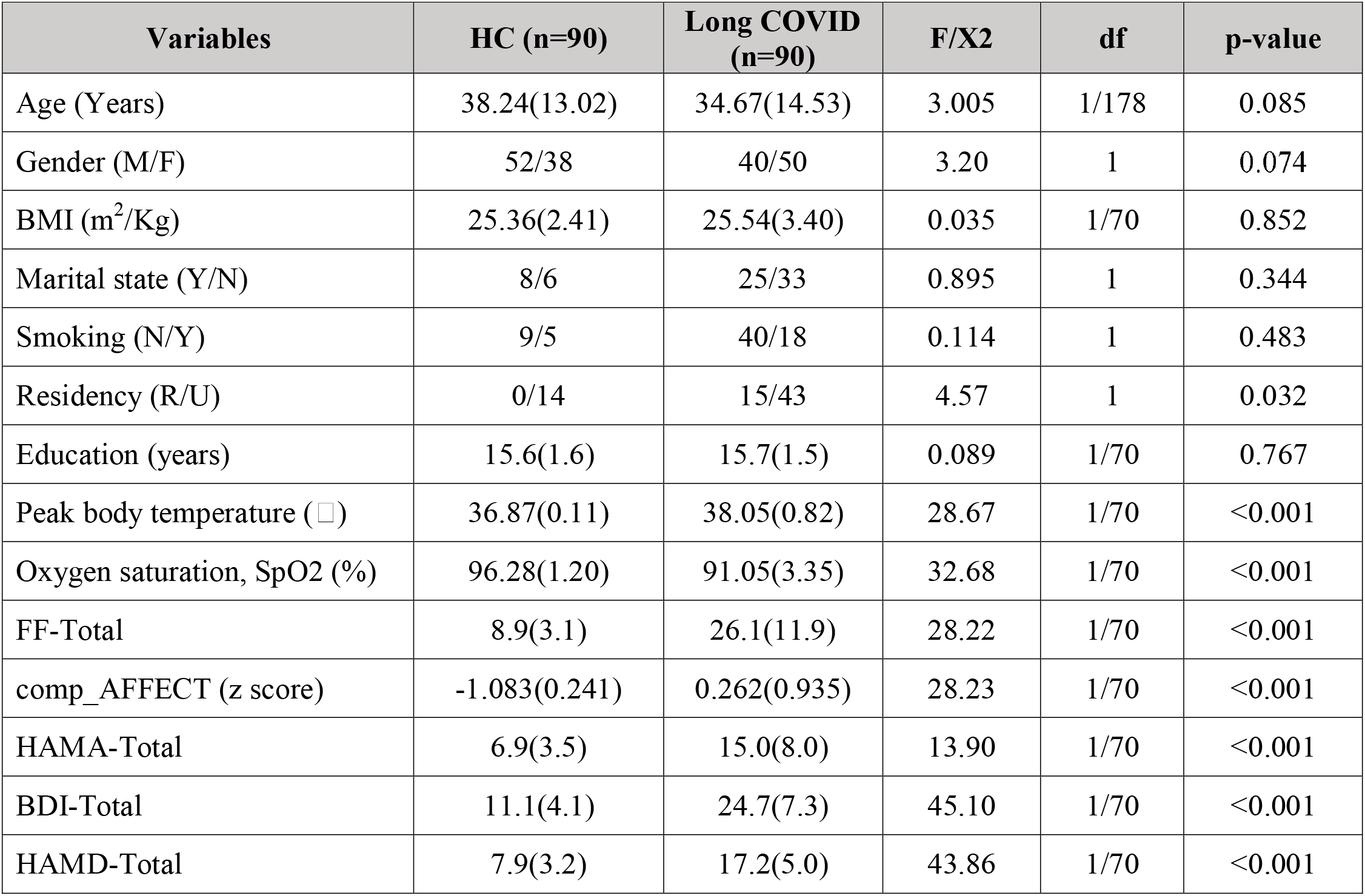
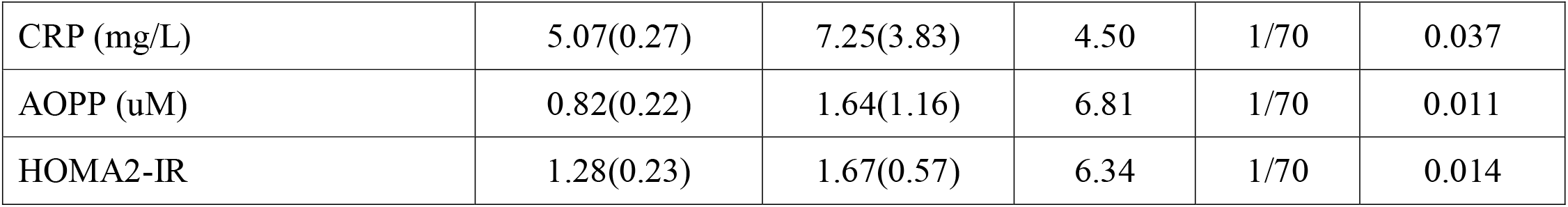
Socio-demographic and clinical data, body temperature, oxygen saturation (SpO2), and psychological rating scales in healthy controls (HC) and Long COVID patients.

PBT was significantly and positively correlated with the HAMD (r=0.698, p<0.001), HAMA (r=0.586, p<0.001), BDI (r=0.675, p<0.001), and FF (r=0.619, p<0.001) scores, CRP (r=0.723, p<0.001), and HOMA2-R (r=0.241, p=0.041). SpO2 was significantly and inversely correlated with the HAMD (r=-0.501, p<0.001), HAMA (r=-0.310, p=0.008), BDI (r=-0.401, p<0.001), and FF (r=-0.597, p<0.001) scores, CRP (r=-0.400, p<0.001), AOPP (r=-0.255, p=0.031), and HOMA2-R (r=-0.245, p=0.038).

### Associations between Long COVID and activated immune responses

To identify the most robust predictors of Long COVID disease, we have executed binary logistic regression analyses in the total study group (n=180), designating Long COVID as the dependent variable (with the control group serving as the reference group) and employing the IgA/IgG/IgM responses to the antigens as independent variables, while adjusting for age, sex, and study site (**Table 2**). Regression #1-3 revealed that IgG and IgM-SARS-CoV-2 were significantly and positively associated with Long COVID, while IgA against SARS-CoV-2 was inversely associated. The effect sizes were (without age, sex, and research site) 0.018, 0.085 and 0.028 for IgA, IgG, and IgM, respectively. There were no significant effects of age, sex and research site (and this for all variables). Regressions #4-6 reveal positive associations between IgG and IgM-HHV-6 and Long COVID, with effect sizes of 0.211 and 0.023, respectively. Regressions #7-9 indicate that both IgG and IgM HHV-6 duTPase are highly significantly associated with Long COVID, with Nagelkerke effect sizes of 0.073 and 0.086, respectively. We found that IgA and IgM directed to activin-A were significantly and positively associated with Long COVID (effect sizes were 0.146 and 0.048, respectively), while IgG-activin-A showed an inverse association (effect size was 0.196). **Figure 1** shows that patients with Long COVID have significantly elevated levels of IgA and IgM towards activin-A as compared to controls.

**Figure 1.**
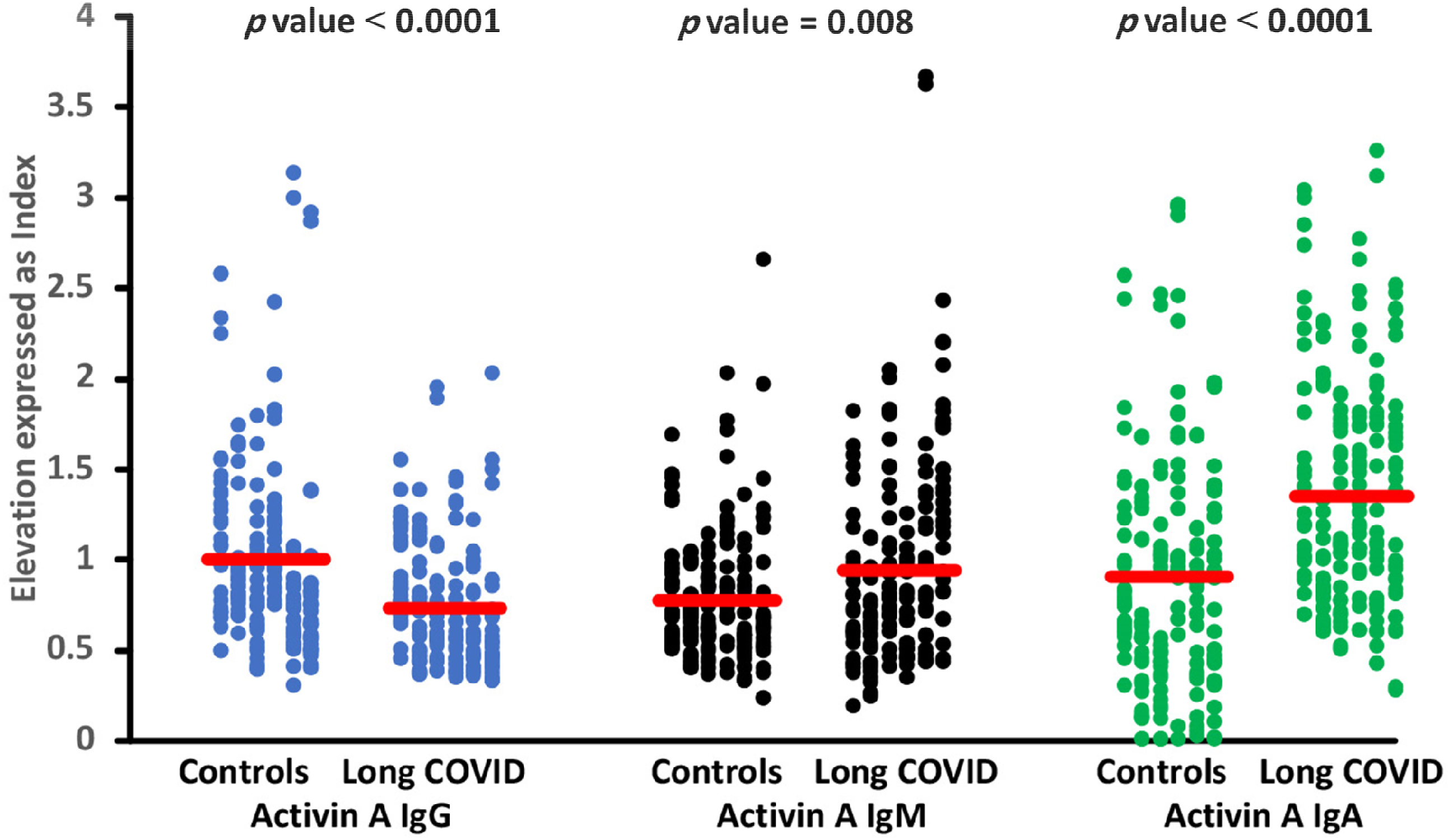
Differences in immunoglobulins IgM, IgA and IgG levels against activin-A between Long COVID patients and controls.

**Table 2:** Results of binary logistic regression analysis with the diagnosis Long COVID as dependent variable (heathy controls as reference group).

| Regression Number | Explanatory Variables | B | SE | Wald | P | OR | 95% CI |
| --- | --- | --- | --- | --- | --- | --- | --- |
| 1 | IgA-SARS-CoV-2 | -0.609 | 0.297 | 4.197 | 0.040 | 0.544 | 0.304 - 0.974 |
| 2 | IgG-SARS-CoV-2 | 1.448 | 0.399 | 13.153 | <0.001 | 4.255 | 1.945 - 9.306 |
| 3 | IgM-SARS-CoV-2 | 0.506 | 0.253 | 4.012 | 0.045 | 1.659 | 1.011 - 2.723 |
| 4 | IgA-HHV-6 | 0.186 | 0.299 | 0.39 | 0.533 | 1.205 | 0.670-2.164 |
| 5 | IgG-HHV-6 | 2.295 | 0.417 | 30.326 | <0.001 | 9.924 | 4.385 - 22.459 |
| 6 | IgM-HHV-6 | 0.475 | 0.23 | 4.271 | 0.039 | 1.608 | 1.025 - 2.523 |
| 7 | IgA-HHV-6-duTPase | -0.114 | 0.287 | 0.158 | 0.691 | 0.892 | 0.509-1.565 |
| 8 | IgG-HHV-6-duTPase | 1.580 | 0.424 | 13.863 | <0.001 | 4.855 | 2.113 - 11.153 |
| 9 | IgM-HHV-6-duTPase | 0.956 | 0.260 | 13.501 | <0.001 | 2.601 | 1.562 - 4.331 |
| 10 | IgA-activin-A | 1.267 | 0.239 | 28.038 | <0.001 | 3.551 | 2.221 - 5.676 |
| 11 | IgG-activin-A | -2.497 | 0.446 | 31.316 | <0.001 | 0.082 | 0.034 - 0.197 |
| 12 | IgM-activin-A | 0.796 | 0.298 | 7.123 | 0.008 | 2.216 | 1.235 - 3.975 |
| 13 | PC_IgA | -0.415 | 0.154 | 7.321 | 0.007 | 0.660 | 0.489-0.892 |
| 14 | IgG-SARS-CoV-2 | 0.726 | 0.199 | 13.32 | <0.001 | 2.067 | 1.400; 3.052 |
|  | IgG-HHV-6 | 0.818 | 0.226 | 13.10 | <0.001 | 2.265 | 1.455; 3.527 |
|  | IgM-HHV-6-duTPase | 0.617 | 0.205 | 9.06 | 0.003 | 1.853 | 1.240; 2.769 |
|  | IgA-activin-A | 1.681 | 0.272 | 38.12 | <0.001 | 5.369 | 3.149; 9.153 |
|  | PC_IgA | -1.786 | 0.276 | 42.01 | <0.001 | 0.168 | 0.098; 0.288 |

OR: Odd's ratio, 95 CI: 95% confidence intervals. Ig: Immunoglobulin, HHV: Human Herpes Virus, duTPase: deoxyuridine 5'-triphosphate nucleotidohydrolase, SARS-CoV-2: severe acute respiratory syndrome coronavirus 2, PC\_IgA: principal component extracted from all IgA values against viral antigens.

There were no significant associations between the Ig responses to citrullinated EBV EBNA2 peptide and EBV-duTPase and Long COVID, except EBV-duTPase which was significantly and negatively associated with Long COVID (Wald=17.29, df=1, p=0.010, Odds ratio=0.232, 95% CI interval: 0.117 – 0.462) with an effect size of 0.080. Interestingly, one PC could be extracted from the IgA responses to IgA responses to SARS-CoV-2, HHV-6, HHV-6-duTPase, citrullinated EBV EBNA2 peptide, and EBV duTPase (labeled PC_IgA, EV=73.6%, KMO=0.845, Χ^2^=1203.72, df=10, p<0.001). **Table 2** shows that this PC was significantly and inversely associated with Long COVID with an effect size of 0.022. There were no significant associations between Long COVID and IgA/IgG/IgM responses to IFN-α2, HSP60, and HSP90.

Regression #14 shows the results of an automatic multivariable logistic regression analysis with Long COVID as outcome variable (and no Long COVID as reference group). Thus, the best prediction of Long COVID (Χ^2^=138.46, df=5, p<0.001) was obtained when PC_IgA (inversely associated) was combined with IgG-SARS-CoV-2, IgG-HHV-6, IgM-HHV-6-duTPase, and IgA-activin-A (all positively associated). The Nagelkerke effect size was 0.533 and 79.8% of all subjects were correctly classified with a sensitivity of 74.6% and specificity of 84.2%.

**Table 3**, neural network model #1 (NN#1) shows the characteristics of a first neural network model discriminating Long COVID from controls. This model was constructed using 2 hidden layers with 7 nodes in hidden layer 1, 5 in hidden layer 2, and 2 units in the output layer. The error term was significantly lower in the testing than in the training sample, whilst the percentage of incorrect classifications was fairly constant between the three samples, indicating that the model was not overtrained. This solution was better than the logistic regression, with a predictive accuracy (computed in the holdout sample) of 80.6% (sensitivity=78.9% and specificity=81.8%) and an area ROC curve of 0.876. The topmost important biomarkers (**Figure 2**) were in descending order of importance: IgA-activin-A, IgG-HHV-6, PC_IgA, IgM-HHV-6-duTPase, and IgG-SARS-CoV-2

**Table 3.** Results of neural networks (NN) with the diagnosis Long COVID or the severity of the Long COVID phenome as output variables and immune variables as input data.

|  | **Models** | **NN#1** | **NN#2** | **NN#3** |
| --- | --- | --- | --- | --- |
| **Input Layer** | Number of units | 9 | 15 | 15 |
| **Hidden layers** | Number of hidden layers | 2 | 2 | 2 |
|  | Activation function | Sigmoid | Sigmoid | Hyperbolic tangent |
|  | Number of units in hidden layer 1 | 7 | 1 | 2 |
|  | Number of units in hidden layer 2 | 5 | 1 | 21 |
| **Output layer** | Dependent variable | Long COVID vs controls | Comp_AFFECT (HAMD+HAMA+BDI) | Total FF |
|  | Number of units | 2 | 1 | 1 |
|  | Activation function | Softmax | Sigmoid | Identity |
| **Training** | Error term | 49.549 (cross-entropy) | 0.679 (sum of squares) | 9.877 (sum of squares) |
|  | % incorrect or relative error | 21.2% | 0.821 | 0.664 |
|  | Prediction (sens, spec) | 74.5%, 82.5% | - | - |
| **Testing** | Sum of Squares error | 30.259 | 0.366 | 0.803 |
|  | % incorrect or relative error | 26.2% | 0.847 | 0.589 |
|  | Prediction: sens, spec | 75.8%, 71.4% | - | - |
|  | AUC ROC | 0.876 | - | - |
| **Holdout** | % incorrect or relative error | 19.4% | 0.950 | 0.764 |
|  | Predictive accuracy: Sens, spec or r value (predicted vs observed) | 78.9%, 81.8% | r=0.632 | 0.709 |

**Figure 2.**
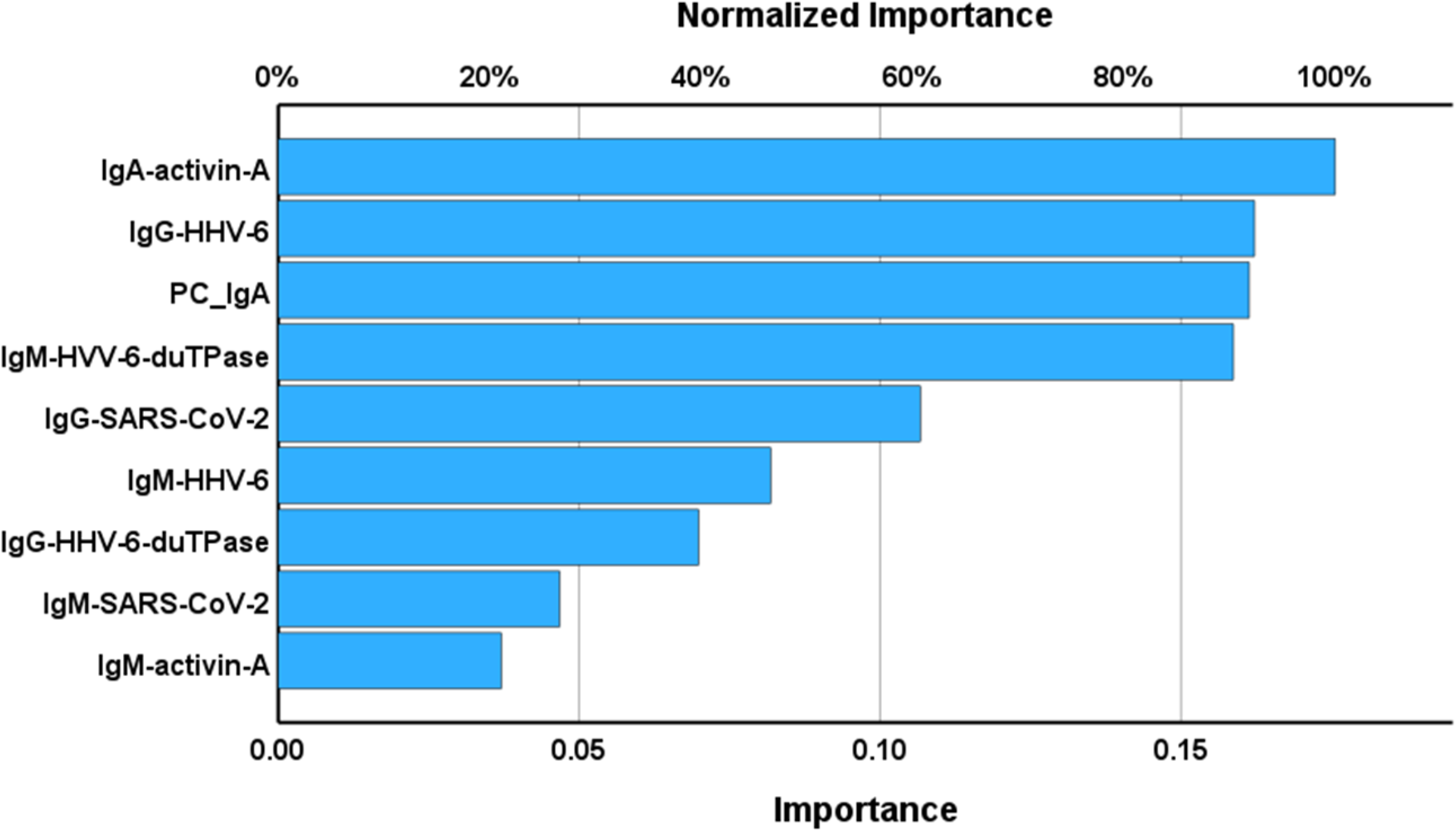
Importance chart of a neural network analysis with Long COVID patients versus controls as output variables. Ig: Immunoglobulin. HHV: Human Herpes Virus, duTPase: deoxyuridine 5′-triphosphate nucleotidohydrolase, SARS-CoV-2: severe acute respiratory syndrome coronavirus 2, PC_IgA: principal component extracted from all IgA values against viral antigens.

There were no significant correlations between the biomarkers, PBT or SpO2, and the IgA/IgG/IgM responses listed in Table 2, except between PBT and IgA-activin-A (r=0.323, p=0.006), but after false discovery p-correction, the latter was no longer significant.

### Results of neural networks with phenome rating scores as output variables

We also analyzed the effects of the abovementioned biomarkers on affective and FF scores. **Figure 3** and **Table 3** depict the features of NN#2, which used affective symptoms (comp_AFFECT) as output variable. This model was trained with 2 hidden layers, each with 1 unit, and with sigmoid as the activation function for the hidden and output layers. During training, the neural network model improved its ability to generalize from the trend, as measured by a decrease in the error term. In addition, the model is not overfitted because the relative error terms were quite similar in the training, testing, and holdout samples. The predicted versus observed r value was 0.632. The relative (normalized) importance of all input variables is depicted in **Figure 3’s** relevance chart. The top predictors that showed the highest predictive value of the model were in descending order: IgM-HHV-6-duTPase, IgG-HHV-6, CRP, education, and IgA-activin-A, followed at a distance by IgG-SARS-CoV-2, IgM-HHV-6, and IgG-HHV-6-duTPase.

**Figure 3.**
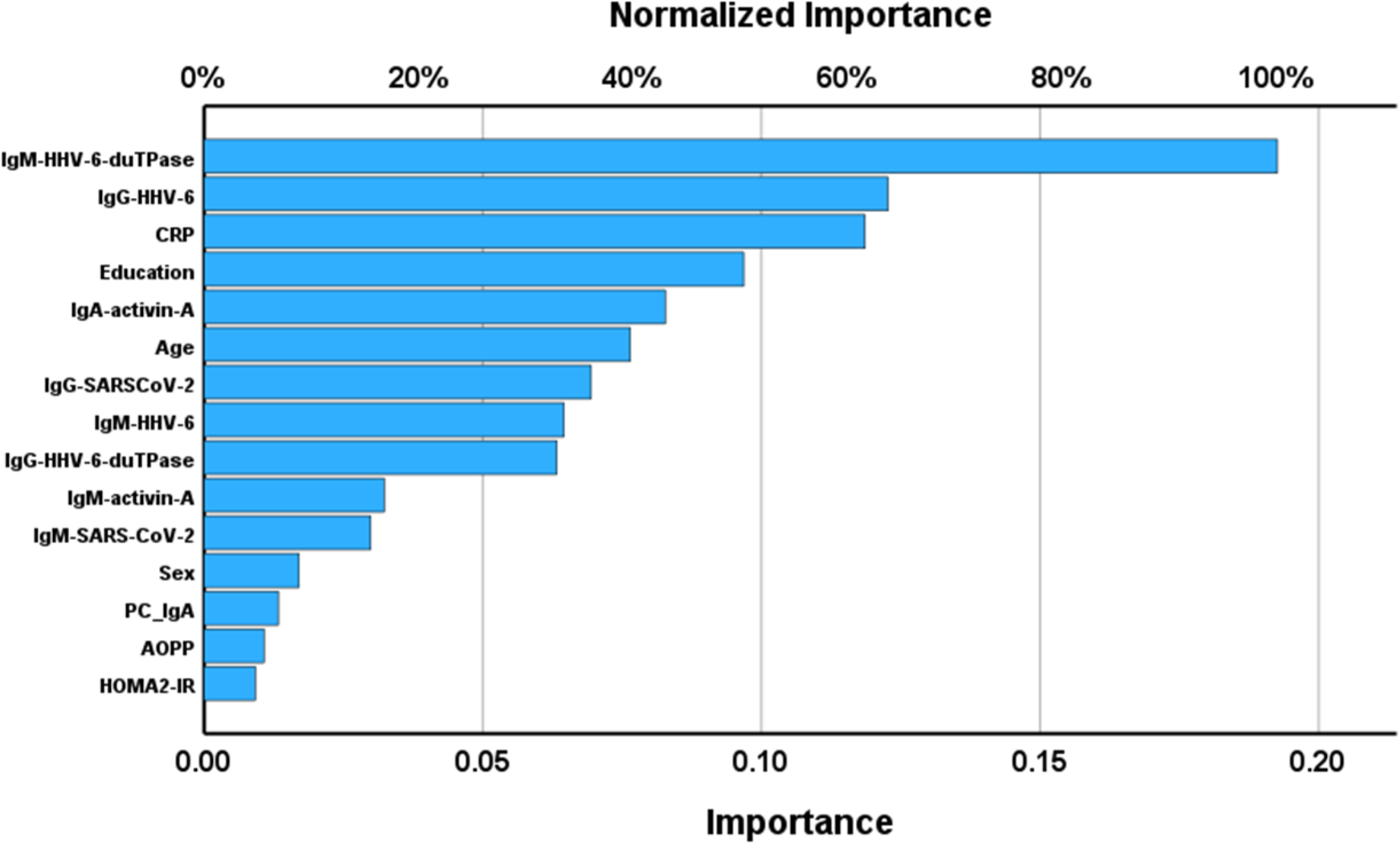
Importance chart of neural network analysis with an affective composite score as dependent variable. Ig: Immunoglobulin. HHV: Human Herpes Virus, duTPase: deoxyuridine 5′-triphosphate nucleotidohydrolase, SARS-CoV-2: severe acute respiratory syndrome coronavirus 2, PC_IgA: principal component extracted from all IgA values against viral antigens, AOPP: advanced oxidation protein products, CRP: C-reactive protein, HOMA2-IR: Homeostatic Model Assessment for Insulin Resistance.

NN#3 used the total FF score as an output variable and was trained using a two layer, architecture. The hyperbolic tangent was used as the activation function during training on this layer, and identity in the output layer. After training, the neural network model could more accurately predict future values by reducing the error term (sum of squares) from 9.877 to 1.213. The training (0.746), testing (0.589), and holdout (0.764) samples had quite similar relative error terms, showing that the model is not overfitted. The predicted versus observed r value was 0.709. The relative (normalized) importance of the input variables is depicted in **Figure 4’s** relevance chart. The top-5 predictors were in descending order of importance: CRP, IgG-HHV-6-duTPase, IgM-activin-A, IgM-SARS-CoV-2, and IgA-activin-A, followed at a distance by IgM-HHV6, AOPP, sex, IgG-SARS-CoV-2 and education. **Figure 5** shows the predicted versus observed value of the NN#3 model.

**Figure 4.**
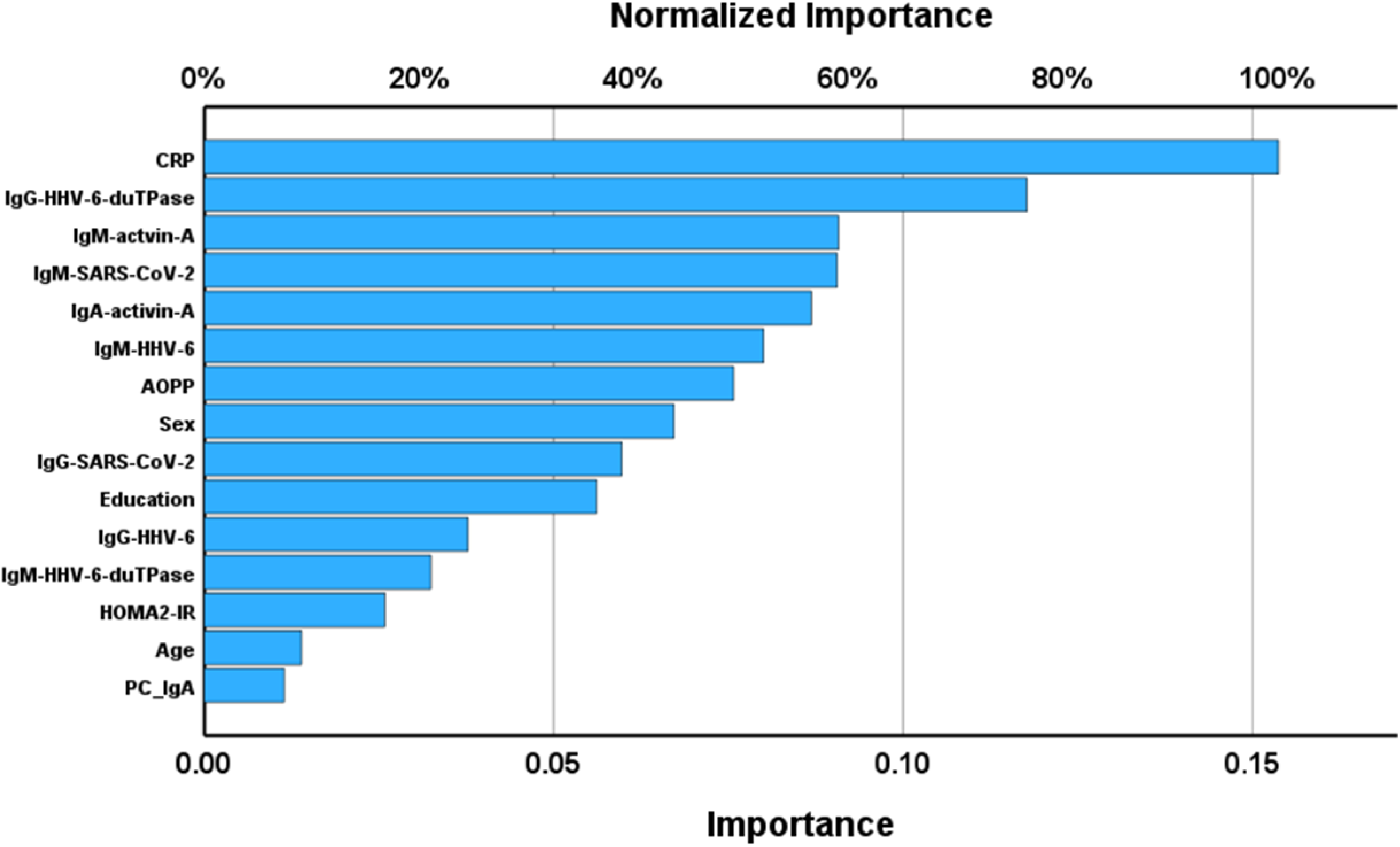
Importance chart of neural network analysis with to total Fatigue-Fibromyalgia (FF) score as dependent variable. Ig: Immunoglobulin. HHV: Human Herpes Virus, duTPase: deoxyuridine 5′-triphosphate nucleotidohydrolase, SARS-CoV-2: severe acute respiratory syndrome coronavirus 2, PC_IgA: principal component extracted from all IgA values against viral antigens, AOPP: advanced oxidation protein products, CRP: C-reactive protein, HOMA2-IR: Homeostatic Model Assessment for Insulin Resistance.

**Figure 5.**
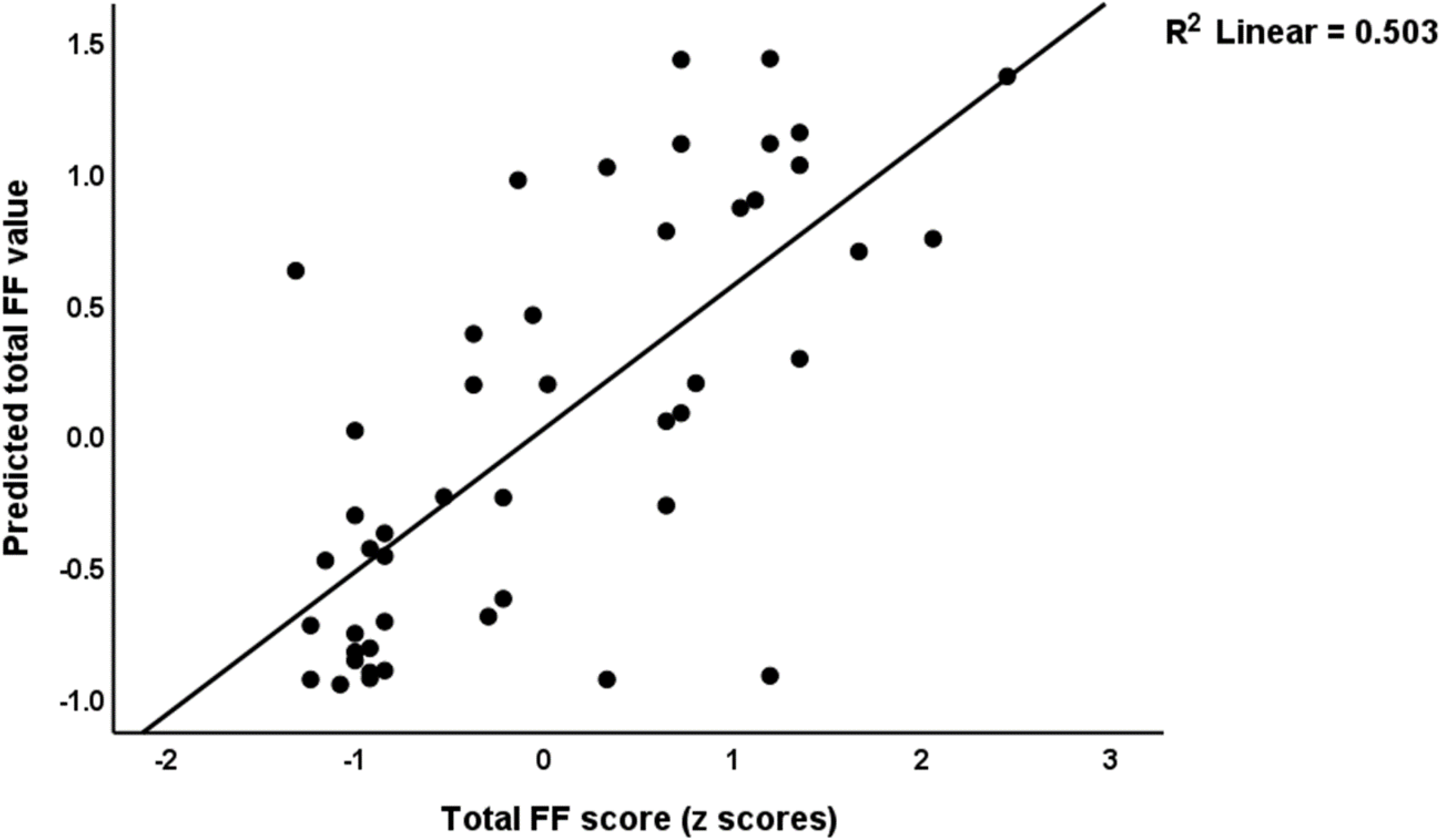
Predictive accuracy of the neuronal network (NN#3) shown in Figure 4 and Table 3; the predicted versus observed value of the Fibro-Fatigue score.

## Discussion

### Long COVID is associated with an immune response

The main finding of this study suggests that Long COVID disease is distinguished from controls by an augmented immune-inflammatory response, as demonstrated by a significant increase in the levels of specific immunoglobulins to viral antigens and self antigens, CRP and AOPP. Prior studies have provided evidence indicating that the activation of immune-inflammatory pathways, characterized by heightened cytokine production, increased CRP levels, the involvement of the NLRP3 inflammasome, oxidative stress pathways, and lowered antioxidant levels play a crucial role in the pathophysiology and persistence of Long COVID disease (Al-Hakeim, Al-Rubaye et al. 2022, Al-Hakeim, Al-Rubaye et al. 2022, Al-Hakeim, Khairi Abed et al. 2023, Almulla, Al-Hakeim et al. 2023, Low, Low et al. 2023). The current investigation additionally demonstrates that Long COVID is characterized by elevated levels of IgA, IgG and/or IgM antibodies targeting HHV-6, HHV-6-duTPase, SARS-CoV-2, and activin-A. These findings extend those of a recent review showing that persistence of inflammation, and immune dysregulation are common patterns leading to or maintaining Long COVID (Vojdani, Vojdani et al. 2023).

Hence, the findings of our study could suggest a potential association between the activated immune and oxidative pathways of Long COVID and increased immunoglobulin levels in SARS-CoV-2, HHV-6, HHV-6-duTPase and activin-A. The latter immune responses to viruses and activin-A may, in theory, play a role in initiating or maintaining the immune-inflammatory response that is characteristic of Long COVID (Vojdani, Vojdani et al. 2023). However, the current study did not find any statistically significant correlations between CRP or AOPP and any of the IgA/IgG/IgM responses. Furthermore, our study did not observe any discernible impacts of PBT or SpO2 on the IgA/IgG/IgM responses during the acute infectious phase. In contrast, we found significant correlations between PBT (positively) and SpO2 (inversely) and CRP, AOPP, and the HOMA2-IR index. As previously mentioned in the introduction, it can be inferred that the intensity of the acute infectious stage is indicative of the subsequent immune and oxidative responses, but not the IgA/IgM/IgG responses to viral antigens and self-antigens measured here.

In agreement with our previous studies (Al-Hakeim, Al-Rubaye et al. 2023), the current study found that the HOMA-2IR index was higher in Long COVID patients than in controls. Nevertheless, no associations were detected between the increased HOMA2-IR index and the IgA/IgM/IgG responses examined in the present study.

### Reactivation of HHV-6 and autoimmune responses to activin-A predict Long COVID

The second important finding from this study is that the clinical diagnosis of Long COVID disease is predicted by elevated levels of IgM/IgG against HHV-6 and its duTPase, and IgG/IgM directed to SARS-CoV-2, as well as IgA/IgM against activin-A, while there are no increased immunoglobulin responses to EBV, IFN-α2 or HSP60/90. Our findings indicate that the pathophysiology of Long COVID is associated with reactivation of HHV-6, SARS-CoV-2 persistence, and autoimmunity to activin-A. A previous review (Vojdani, Vojdani et al. 2023) summarized that persistent SARS-CoV-2 infection, HHV-6 reactivation, and associated processes during Long COVID contribute to the pathophysiology of Long COVID. Nevertheless, it is unclear whether latent viruses amplify the immunological response that is important in initiating Long COVID or whether they directly contribute to the disease.

The findings presented in this study extend those of prior research which documented reactivation of HHV-6 by detecting DNA of the virus in 25% of the individuals with Long COVID (Zubchenko, Kril et al. 2022). The underlying mechanism may be explained by the interference with the host’s type I interferon response, which may be disrupted by autoantibodies (Acharya, Liu et al. 2020), resulting in a compromised ability to control latent pathogens (Rojas, Rodríguez et al. 2022, Vojdani, Vojdani et al. 2023). However, the current study did not find any significant alterations in the IgA/IgG/IgM-mediated responses to IFN-α2.

Furthermore, SARS-COV-2 persistence may be a factor in some patients since IgM to SARS-CoV-2 is connected to the Long COVID diagnosis, albeit with a smaller impact size (see neural network data). Previously, Su et al. (Su, Yuan et al. 2022) reported that many individuals with Long COVID had antibodies against the receptor binding domain of the SARS-CoV-2 spike protein. This indicates the persistence of SARS-CoV-2 virus even after months of full recovery. As a result, the ongoing presence of SARS-CoV-2 and its secreted superantigens, which are known to trigger polyclonal T-cell activation, may cause immune activation, including dendritic cell activation, and apoptotic processes in the hosts’ cells. Consequently, this process may lead to autoimmune responses seen in Long COVID patients (Jacobs 2021, Vojdani, Vojdani et al. 2023).

Previous reports found EBV reactivation in Long COVID, as indicated by an increase in VCA IgM and EA-D IgG levels (Gold, Okyay et al. 2021). Only a few patients showed positive EBV viremia from nasal swabs 2-3 months after recovery (Su, Yuan et al. 2022). Likewise, in the current investigation, we were unable to demonstrate elevated immunoglobulin responses to EBV antigens in Long COVID.

Intriguingly, our analysis found that the Long COVID diagnosis was inversely correlated with the first component extracted from all IgA levels (labeled: PC_IgA) to viral antigens (SARS-CoV-2, HHV-6, EBV, and duTPase from HHV-6 and EBV). IgA is effective against certain viruses, including SARS-CoV-2 (Quinti, Mortari et al. 2021, Sterlin, Mathian et al. 2021). The severity of COVID-19 and prolonged viral shedding may be aggravated by decreased anti-SARS-Cov-2 (and other viruses) IgA and secretory IgA (sIgA) (Quinti, Mortari et al. 2021). As a result, we may posit that Long COVID may develop, especially in subjects with decreased IgA against viral antigens or maybe abnormalities in IgA class switching.

Importantly, our neural network analysis shows that the diagnosis of Long COVID is well classified with a predictive accuracy of 80.6% (sensitivity of 78.9% and specificity of 81.8%); the top discriminatory variables are IgA-activin-A, IgG-HHV-6, IgM-HVV-6 duTPase, IgG-SARS-CoV-2, and IgM-HHV-6 (all positively) and PC_IgA (inversely). These results of our univariate and multivariate analyses show that Long COVID is to a large extent the consequence of reactivation of HVV-6, autoimmune responses to activin-A, activated immune and oxidative stress pathways, and to a lesser extent SARS-CoV-2 persistence.

### IgA/IgG/IgM responses to antigens predict the severity of the Long COVID phenome

The third major finding of this study is that a combination of different immunoglobulins to viral and self-antigens and inflammatory biomarkers predicts the severity of the affective symptoms and CFS due to Long COVID; the top predictors were (in descending order): CRP, IgA-Activin-A, IgG-HHV-6-duTPase, IgG-HHV-6, IgM-HHV-6-duTPase, and IgM-HHV-6. These findings indicate that reactivation of HHV-6, IgA-mediated responses to activin-A, and activated immune-inflammatory pathways predict the severity of the Long COVID phenome.

Long-term COVID patients display symptoms comparable to those observed in myalgic encephalomyelitis (ME)/CFS, a condition characterized by severe fatigue, musculoskeletal pain, and post-exercise malaise, as well as affective symptoms and neurocognitive deficits (Morris and Maes 2013). A previous longitudinal study demonstrated that, in Long COVID, fatigue was associated with EBV viremia and that in acute COVID-19 memory impairment was associated with EBV viremia and SARS-CoV-2 RNAemia (Su, Yuan et al. 2022). There is evidence that ME/CFS and affective disorders are associated with activated immune-inflammatory and oxidative pathways, while ME/CFS also has strong associations with a variety of autoimmune processes (Twisk and Maes 2009, Morris and Maes 2013; 2017, Komaroff and Bateman 2021, Kedor, Freitag et al. 2022). In addition, ME/CFS is frequently precipitated by viral infections, such as HHV-6, or repeated exposure to pathogens, suggesting that infectious agents such as HHV-6 play a role in both ME/CFS and Long COVID (Maes, Twisk et al. 2012, Morris and Maes 2013, Rasa, Nora-Krukle et al. 2018). Importantly, the substantial increase in virus-specific HHV-6 antibodies in saliva, is especially pronounced in patients with ME/CFS (Apostolou, Rizwan et al. 2022). These findings emphasize similarities in antiviral profiles against latent viruses in ME/CFS and Long COVID (Apostolou, Rizwan et al. 2022).

Our findings suggest that activin-A abnormalities play a significant role in the severity of affective symptoms and CFS due to Long COVID. Indeed, elevated IgA mediated autoimmunity against activin-A is the most significant predictor of the diagnosis Long COVID, while IgA/IgM responses to activin-A are major predictors of the severity of the affective phenome and CFS symptoms. As described in the Introduction, activin-A, as a member of the TGF-β family, is a crucial component of immune function. Inflammatory cytokines, Toll-like receptor ligands, and oxidative stress stimulate the production and release of activins (Gravelsina, Nora-Krukle et al. 2021). Activin-A regulates the growth and differentiation of various immune cell types and immune responses and plays a role in the activin/follistatin-axis, which is significantly disrupted in COVID-19 cases and is linked to higher mortality rates (Bilezikjian, Blount et al. 2006, Megan, Qian et al. 2021, Synolaki, Papadopoulos et al. 2021). Activin-A has been shown to inhibit virus replication in multiple infected cell lines (Lidbury, Kita et al. 2017) and inhibits virus replication in the A549 lung epithelial cell line following Zika virus infection, either alone or in combination with IFN-α (Eddowes, Al-Hourani et al. 2019). Activin-A plays a dual function in autoimmune diseases, exhibiting both pro-inflammatory and anti-inflammatory properties. Specifically, it regulates the inflammatory responses associated with pathogenic T helper 1 and T helper 17 cells in the central nervous system (CNS) (Morianos, Papadopoulou et al. 2019).

Importantly, aberrations in activin are known to influence depressive and anxiety related behaviors (Dow, Russell et al. 2005, Ageta, Murayama et al. 2008), and play a role in the pathophysiology of ME/CFS (Lidbury, Kita et al. 2017). Additionally, activin is involved in cognitive functioning (Zheng, Link et al. 2017). The expression of activin-A increases in response to neuronal activity (Andreasson and Worley 1995, Inokuchi, Kato et al. 1996) and may protect neurons from ischemic injury (Tretter, Hertel et al. 2000). It influences dendritic spine morphology, a critical factor for synaptic plasticity in the hippocampus (Fukazawa, Saitoh et al. 2003, Shoji-Kasai, Ageta et al. 2007). Recent research by Dow and colleagues (Dow, Russell et al. 2005) demonstrated that administration of activin into the hippocampus produced similar effects to antidepressants. In addition, the concentration of activin in the hippocampus appears to influence behaviors associated with depression and anxiety. Therefore, activin may be a novel modulator of affective symptoms (Ageta, Murayama et al. 2008). Activin B, which shares 65% sequence homology with Activin-A (Massagué 1987), has also been identified as a novel biomarker for CFS. Lidbury, Kita, and colleagues (2017) suggest that analyzing levels of activin and its binding protein, follistatin, could be a valuable tool for distinguishing CFS from other fatigue-related disorders (Lidbury, Kita et al. 2017). Activin may therefore play an important role in the pathophysiology of affective and CFS symptoms due to Long COVID.

## Limitations

While interpreting the current data, it is imperative to acknowledge certain limitations. Specifically, an examination of immunoglobulins against a broader spectrum of latent viruses - including CMV, HHV-7, Herpes Simplex Virus types 1 and 2, Human Papillomavirus, and Adenovirus – will enrich future studies. More studies are required to investigate the associations between immune-inflammatory pathways, including the NLRP3 inflammasome, and oxidative and nitrosative stress, including hypernitrosylation, and SARS-CoV-2 persistence, latent virus reactivation, and autoimmune reactivity in Long COVID disease.

## Conclusions

Long COVID patients demonstrate elevated levels of CRP, AOPP, HOMA2-IR, IgG/IgM-SARS-CoV-2, IgG/IgM-HVV-6 and HVV-6-duTPase, and IgA/IgM-activin-A. Using the combination of these data in neural networks, we obtained adequate predictive accuracy, predicting Long COVID. The severity of the Long COVID phenome, which includes depression, anxiety, and CFS symptoms, was predicted by immune and oxidative biomarkers, with CRP, IgM/IgG HHV-6, IgM/IgG-HHV-6-duTPase, and IgM/IgA-activin-A as top predictors. These data show that interactions among these different biomarkers are associated with the development or maintenance of Long COVID. The results show that reactivation of HHV-6, persistence of SARS-CoV-2, and autoimmune reactions to activin-A, in conjunction with activated immune-oxidative pathways and increased insulin resistance, play a significant role in the pathophysiology of Long COVID.

## Data Availability

AA and AV oversaw taking blood samples and managing other patient-related duties. AV quantified the biomarkers in the blood serum. MM conducted the study's statistical analysis. The work is written and edited by AA, MM, AV and HAH. All authors have read and approved the final manuscript.

## Acknowledgments

The authors would like to express their gratitude to all individuals who helped with the data collection at the Imam Sajjad Hospital, Hassan Halos Al-Hatmy Hospital for Transmitted Diseases, Middle Euphrates Center for Cancer, Al-Najaf Teaching Hospital, and Al-Sader Medical City of Najaf.

## Ethical approval and consent to participate

The College of Medical Technology Ethics Committee approved this study at the Islamic University of Najaf, Iraq (Document No. 34/2023). The research was conducted strictly per local, international, and Iraqi ethical and privacy laws. Written informed consent was obtained from patients and controls.

## Declaration of interest

The authors declare no conflicts of interest.

## Funding

The C2F program at Chulalongkorn University in Thailand, grant number 64.310/436/2565 to AFA, the Thailand Science Research, and Innovation Fund at Chulalongkorn University (HEA663000016), and a Sompoch Endowment Fund (Faculty of Medicine) MDCU (RA66/016) to MM all provided funding for the project.

## Availability of data

The corresponding author (MM) will make the SPSS file used in the current study available upon receipt of an appropriate request and once the author has fully exploited the data.

## Notes

### Competing Interest Statement

The authors have declared no competing interest.

## References

1. Acharya, D., G. Liu and M. U. Gack (2020). “Dysregulation of type I interferon responses in COVID-19.” Nature Reviews Immunology 20(7): 397–398.

2. Ageta, H., A. Murayama, R. Migishima, S. Kida, K. Tsuchida, M. Yokoyama and K. Inokuchi (2008). “Activin in the brain modulates anxiety-related behavior and adult neurogenesis.” PLoS One 3(4): e1869.

3. Al-Hadrawi, D. S., H. T. Al-Rubaye, A. F. Almulla, H. K. Al-Hakeim and M. Maes (2022). “Lowered oxygen saturation and increased body temperature in acute COVID-19 largely predict chronic fatigue syndrome and affective symptoms due to Long COVID: A precision nomothetic approach.” Acta Neuropsychiatrica: 1–12.

4. Al-Hakeim, H. K., H. T. Al-Rubaye, D. S. Al-Hadrawi, A. F. Almulla and M. Maes (2022). “Long-COVID post-viral chronic fatigue and affective symptoms are associated with oxidative damage, lowered antioxidant defenses and inflammation: a proof of concept and mechanism study.” Molecular Psychiatry.

5. Al-Hakeim, H. K., H. T. Al-Rubaye, A. F. Almulla, D. S. Al-Hadrawi and M. Maes (2022). “Chronic fatigue, depression and anxiety symptoms in Long COVID are strongly predicted by neuroimmune and neuro-oxidative pathways which are caused by the inflammation during acute infection.” medRxiv: 2022.2006.2029.22277056.

6. Al-Hakeim, H. K., H. T. Al-Rubaye, A. S. Jubran, A. F. Almulla, S. R. Moustafa and M. Maes (2023). “Increased insulin resistance due to Long COVID is associated with depressive symptoms and partly predicted by the inflammatory response during acute infection.” Braz J Psychiatry.

7. Al-Hakeim, H. K., A. Khairi Abed, S. Rouf Moustafa, A. F. Almulla and M. Maes (2023). “Tryptophan catabolites, inflammation, and insulin resistance as determinants of chronic fatigue syndrome and affective symptoms in long COVID.” Frontiers in Molecular Neuroscience 16.

8. Al-Jassas, H. K., H. K. Al-Hakeim and M. Maes (2022). “Intersections between pneumonia, lowered oxygen saturation percentage and immune activation mediate depression, anxiety, and chronic fatigue syndrome-like symptoms due to COVID-19: A nomothetic network approach.” J Affect Disord 297: 233–245.

9. Almulla, A. F., H. K. Al-Hakeim and M. Maes (2023). “Chronic fatigue and affective symptoms in acute and long COVID are attributable to immune-inflammatory pathways.” Psychiatry Clin Neurosci 77(2): 125–126.

10. Anderson, G., A. F. Almulla, R. J. Reiter and M. Maes (2023) “Redefining Autoimmune Disorders’ Pathoetiology: Implications for Mood and Psychotic Disorders’ Association with Neurodegenerative and Classical Autoimmune Disorders.” Cells 12 DOI: 10.3390/cells12091237.

11. Andreasson, K. and P. F. Worley (1995). “Induction of beta-A activin expression by synaptic activity and during neocortical development.” Neuroscience 69(3): 781–796.

12. Apostolou, E., M. Rizwan, P. Moustardas, P. Sjögren, B. C. Bertilson, B. Bragée, O. Polo and A. Rosén (2022). “Saliva antibody-fingerprint of reactivated latent viruses after mild/asymptomatic COVID-19 is unique in patients with myalgic encephalomyelitis/chronic fatigue syndrome.” Frontiers in Immunology 13: 6407.

13. Balc’h, L., K. Pinceaux, C. Pronier, P. Seguin, J.-M. Tadié and F. Reizine (2020). “Herpes simplex virus and cytomegalovirus reactivations among severe COVID-19 patients.” Critical Care 24(1): 1–3.

14. Bilezikjian, L. M., A. L. Blount, C. J. Donaldson and W. W. Vale (2006). “Pituitary actions of ligands of the TGF-β family: activins and inhibins.” Reproduction 132(2): 207–215.

15. Chen, B., B. Julg, S. Mohandas, S. B. Bradfute and R. M. P. T. Force (2023). “Viral persistence, reactivation, and mechanisms of long COVID.” eLife 12: e86015.

16. Cheng, M. H., S. Zhang, R. A. Porritt, M. Noval Rivas, L. Paschold, E. Willscher, M. Binder, M. Arditi and I. Bahar (2020). “Superantigenic character of an insert unique to SARS-CoV-2 spike supported by skewed TCR repertoire in patients with hyperinflammation.” Proceedings of the National Academy of Sciences 117(41): 25254–25262.

17. Cheung, C. C. L., D. Goh, X. Lim, T. Z. Tien, J. C. T. Lim, J. N. Lee, B. Tan, Z. E. A. Tay, W. Y. Wan, E. X. Chen, S. N. Nerurkar, S. Loong, P. C. Cheow, C. Y. Chan, Y. X. Koh, T. T. Tan, S. Kalimuddin, W. M. D. Tai, J. L. Ng, J. G. Low, J. Yeong and K. H. Lim (2022). “Residual SARS-CoV-2 viral antigens detected in GI and hepatic tissues from five recovered patients with COVID-19.” Gut 71(1): 226–229.

18. Cox, B. S., K. Alharshawi, I. Mena-Palomo, W. P. Lafuse and M. E. Ariza (2022). “EBV/HHV-6A dUTPases contribute to myalgic encephalomyelitis/chronic fatigue syndrome pathophysiology by enhancing TFH cell differentiation and extrafollicular activities.” JCI Insight 7(11).

19. Dow, A. L., D. S. Russell and R. S. Duman (2005). “Regulation of activin mRNA and Smad2 phosphorylation by antidepressant treatment in the rat brain: effects in behavioral models.” J Neurosci 25(20): 4908–4916.

20. Eddowes, L. A., K. Al-Hourani, N. Ramamurthy, J. Frankish, H. T. Baddock, C. Sandor, J. D. Ryan, D. N. Fusco, J. Arezes and E. Giannoulatou (2019). “Antiviral activity of bone morphogenetic proteins and activins.” Nature microbiology 4(2): 339–351.

21. Fukazawa, Y., Y. Saitoh, F. Ozawa, Y. Ohta, K. Mizuno and K. Inokuchi (2003). “Hippocampal LTP is accompanied by enhanced F-actin content within the dendritic spine that is essential for late LTP maintenance in vivo.” Neuron 38(3): 447–460.

22. Gerwyn, M. and M. Maes (2017). “Mechanisms Explaining Muscle Fatigue and Muscle Pain in Patients with Myalgic Encephalomyelitis/Chronic Fatigue Syndrome (ME/CFS): a Review of Recent Findings.” Curr Rheumatol Rep 19(1): 1.

23. Gold, J. E., R. A. Okyay, W. E. Licht and D. J. Hurley (2021) “Investigation of Long COVID Prevalence and Its Relationship to Epstein-Barr Virus Reactivation.” Pathogens 10 DOI: 10.3390/pathogens10060763.

24. Gravelsina, S., Z. Nora-Krukle, A. Vilmane, S. Svirskis, K. Vecvagare, A. Krumina and M. Murovska (2021). “Potential of Activin B as a Clinical Biomarker in Myalgic Encephalomyelitis/Chronic Fatigue Syndrome (ME/CFS).” Biomolecules 11(8).

25. Groff, D., A. Sun, A. E. Ssentongo, D. M. Ba, N. Parsons, G. R. Poudel, A. Lekoubou, J. S. Oh, J. E. Ericson, P. Ssentongo and V. M. Chinchilli (2021). “Short-term and Long term Rates of Postacute Sequelae of SARS-CoV-2 Infection: A Systematic Review.” JAMA Netw Open 4(10): e2128568.

26. Hamilton, M. (1959). “The assessment of anxiety states by rating.” Br J Med Psychol 32(1): 50–55.

27. Hamilton, M. (1960). “A rating scale for depression.” Journal of neurology, neurosurgery, and psychiatry 23(1): 56–62.

28. Hautzinger, M. K. F. K. h. C. B. A. T. S. R. A. B. G. K. (2009). Beck Depressions-Inventar : BDI-II ; Revision ; Manual. Frankfurt am Main, Pearson.

29. Hu, C., J. Yang, Z. Qi, H. Wu, B. Wang, F. Zou, H. Mei, J. Liu, W. Wang and Q. Liu (2022). “Heat shock proteins: Biological functions, pathological roles, and therapeutic opportunities.” MedComm (2020) 3(3): e161.

30. Inokuchi, K., A. Kato, K. Hiraia, F. Hishinuma, M. Inoue and F. Ozawa (1996). “Increase in activin beta A mRNA in rat hippocampus during long-term potentiation.” FEBS Lett 382(1-2): 48–52.

31. Jacobs, J. J. (2021). “Persistent SARS-2 infections contribute to long COVID-19.” Medical hypotheses 149: 110538.

32. Jon, K., W. Jamie, J. Jillian, L. Peiwen, M. D. Rahul, R. G. Jeff, T. Alexandra, T. Laura, A. M. Amyn, K. Kathy, G. Kerrie, M. Valter Silva, P.-H. Mario, M. Tianyang, B. Bornali, T. Takehiro, L. Carolina, S. Julio, M. Dayna, B. Erica, T.-M. Jenna, D. Yile, P. Emily, A. Koray, J. T. Tiffany, X. Lan, Y. Inci, M. K. Harlan, S. John, M. Ruslan, B. O. Saad, D. David van, M. R. Aaron, P. David and I. Akiko (2022). “Distinguishing features of Long COVID identified through immune profiling.” medRxiv: 2022.2008.2009.22278592.

33. Kedor, C., H. Freitag, L. Meyer-Arndt, K. Wittke, L. G. Hanitsch, T. Zoller, F. Steinbeis, M. Haffke, G. Rudolf, B. Heidecker, T. Bobbert, J. Spranger, H.-D. Volk, C. Skurk, F. Konietschke, F. Paul, U. Behrends, J. Bellmann-Strobl and C. Scheibenbogen (2022). “A prospective observational study of post-COVID-19 chronic fatigue syndrome following the first pandemic wave in Germany and biomarkers associated with symptom severity.” Nature Communications 13(1): 5104.

34. Komaroff, A. L. and L. Bateman (2021). “Will COVID-19 lead to myalgic encephalomyelitis/chronic fatigue syndrome?” Frontiers in Medicine 7: 1132.

35. Lehner, G. F., S. J. Klein, H. Zoller, A. Peer, R. Bellmann and M. Joannidis (2020). “Correlation of interleukin-6 with Epstein–Barr virus levels in COVID-19.” Critical Care 24: 1–3.

36. Li, L., S. Li, Y. Pan, L. Qin, S. Yang, D. Tan, Y. Hu, M. D. Knoll, X. Wang, L. Wang and Q. Wang (2021). “An Immunocompetent Patient with High Neutralizing Antibody Titers Who Shed COVID-19 Virus for 169 days - China, 2020.” China CDC Wkly 3(32): 688–691.

37. Lidbury, B. A., B. Kita, D. P. Lewis, S. Hayward, H. Ludlow, M. P. Hedger and D. M. de Kretser (2017). “Activin B is a novel biomarker for chronic fatigue syndrome/myalgic encephalomyelitis (CFS/ME) diagnosis: a cross sectional study.” J Transl Med 15(1): 60.

38. Low, R. N., R. J. Low and A. Akrami (2023). “A review of cytokine-based pathophysiology of Long COVID symptoms.” Frontiers in Medicine 10.

39. Maes, M. (2022). “Precision Nomothetic Medicine in Depression Research: A New Depression Model, and New Endophenotype Classes and Pathway Phenotypes, and A Digital Self.” J Pers Med 12(3).

40. Maes, M., F. N. Twisk, M. Kubera and K. Ringel (2012). “Evidence for inflammation and activation of cell-mediated immunity in Myalgic Encephalomyelitis/Chronic Fatigue Syndrome (ME/CFS): increased interleukin-1, tumor necrosis factor-α, PMN-elastase, lysozyme and neopterin.” J Affect Disord 136(3): 933–939.

41. Maes, M. H. and D. Stoyanov (2022). “False dogmas in mood disorders research: Towards a nomothetic network approach.” World J Psychiatry 12(5): 651–667.

42. Massagué, J. (1987). “The TGF-beta family of growth and differentiation factors.” Cell 49(4): 437–438.

43. Megan, M., Z. Qian, X. Jianing, P. Li, W. Matthew, J. E. Peter, F. W. Matthew, S. Tea, C. H. Sara, B. Anita, G. M. Lori, A. K. Christos and J. G. David (2021). “Activin A correlates with the worst outcomes in COVID-19 patients, and can be induced by cytokines via the IKK/NF-kappa B pathway.” bioRxiv: 2021.2002.2004.429815.

44. Morianos, I., G. Papadopoulou, M. Semitekolou and G. Xanthou (2019). “Activin-A in the regulation of immunity in health and disease.” Journal of Autoimmunity 104: 102314.

45. Morris, G. and M. Maes (2013). “Myalgic encephalomyelitis/chronic fatigue syndrome and encephalomyelitis disseminata/multiple sclerosis show remarkable levels of similarity in phenomenology and neuroimmune characteristics.” BMC Medicine 11(1): 205.

46. Morris, G. and M. Maes (2013). “A neuro-immune model of Myalgic Encephalomyelitis/Chronic fatigue syndrome.” Metab Brain Dis 28(4): 523–540.

47. Morris, G. and M. Maes (2014). “Mitochondrial dysfunctions in myalgic encephalomyelitis/chronic fatigue syndrome explained by activated immuno inflammatory, oxidative and nitrosative stress pathways.” Metab Brain Dis 29(1): 19–36.

48. Natarajan, A., S. Zlitni, E. F. Brooks, S. E. Vance, A. Dahlen, H. Hedlin, R. M. Park, A. Han, D. T. Schmidtke, R. Verma, K. B. Jacobson, J. Parsonnet, H. F. Bonilla, U. Singh, B. A. Pinsky, J. R. Andrews, P. Jagannathan and A. S. Bhatt (2022). “Gastrointestinal symptoms and fecal shedding of SARS-CoV-2 RNA suggest prolonged gastrointestinal infection.” Med 3(6): 371–387.e379.

49. Proal, A. D. and M. B. VanElzakker (2021). “Long COVID or Post-acute Sequelae of COVID-19 (PASC): An Overview of Biological Factors That May Contribute to Persistent Symptoms.” Front Microbiol 12: 698169.

50. Quinti, I., E. P. Mortari, A. Fernandez Salinas, C. Milito and R. Carsetti (2021). “IgA Antibodies and IgA Deficiency in SARS-CoV-2 Infection.” Front Cell Infect Microbiol 11: 655896.

51. Rahmani, A., G. Dini, V. Leso, A. Montecucco, B. Kusznir Vitturi, I. Iavicoli and P. Durando (2022). “Duration of SARS-CoV-2 shedding and infectivity in the working age population: a systematic review and meta-analysis.” Med Lav 113(2): e2022014.

52. Rasa, S., Z. Nora-Krukle, N. Henning, E. Eliassen, E. Shikova, T. Harrer, C. Scheibenbogen, M. Murovska and B. K. Prusty (2018). “Chronic viral infections in myalgic encephalomyelitis/chronic fatigue syndrome (ME/CFS).” Journal of Translational Medicine 16(1): 1–25.

53. Renaud-Charest, O., L. M. W. Lui, S. Eskander, F. Ceban, R. Ho, J. D. Di Vincenzo, J. D. Rosenblat, Y. Lee, M. Subramaniapillai and R. S. McIntyre (2021). “Onset and frequency of depression in post-COVID-19 syndrome: A systematic review.” Journal of psychiatric research 144: 129–137.

54. Rojas, M., Y. Rodríguez, Y. Acosta-Ampudia, D. M. Monsalve, C. Zhu, Q. Z. Li, C. Ramírez-Santana and J. M. Anaya (2022). “Autoimmunity is a hallmark of post-COVID syndrome.” J Transl Med 20(1): 129.

55. Sagulkoo, P., K. Plaimas, A. Suratanee, A. N. Colado Simão, E. M. Vissoci Reiche and M. Maes (2022). “Immunopathogenesis and Immunogenetic Variants in COVID-19.” Curr Pharm Des 28(22): 1780–1797.

56. Sandler, C. X., V. B. B. Wyller, R. Moss-Morris, D. Buchwald, E. Crawley, J. Hautvast, B. Z. Katz, H. Knoop, P. Little, R. Taylor, K.-A. Wensaas and A. R. Lloyd (2021). “Long COVID and Post-infective Fatigue Syndrome: A Review.” Open forum infectious diseases 8(10): ofab440–ofab440.

57. Shoji-Kasai, Y., H. Ageta, Y. Hasegawa, K. Tsuchida, H. Sugino and K. Inokuchi (2007). “Activin increases the number of synaptic contacts and the length of dendritic spine necks by modulating spinal actin dynamics.” J Cell Sci 120(Pt 21): 3830–3837.

58. Sterlin, D., A. Mathian, M. Miyara, A. Mohr, F. Anna, L. Claër, P. Quentric, J. Fadlallah, H. Devilliers, P. Ghillani, C. Gunn, R. Hockett, S. Mudumba, A. Guihot, C. E. Luyt, J. Mayaux, A. Beurton, S. Fourati, T. Bruel, O. Schwartz, J. M. Lacorte, H. Yssel, C. Parizot, K. Dorgham, P. Charneau, Z. Amoura and G. Gorochov (2021). “IgA dominates the early neutralizing antibody response to SARS-CoV-2.” Sci Transl Med 13(577).

59. Su, Y., D. Yuan, D. G. Chen, R. H. Ng, K. Wang, J. Choi, S. Li, S. Hong, R. Zhang, J. Xie, S. A. Kornilov, K. Scherler, A. J. Pavlovitch-Bedzyk, S. Dong, C. Lausted, I. Lee, S. Fallen, C. L. Dai, P. Baloni, B. Smith, V. R. Duvvuri, K. G. Anderson, J. Li, F. Yang, C. J. Duncombe, D. J. McCulloch, C. Rostomily, P. Troisch, J. Zhou, S. Mackay, Q. DeGottardi, D. H. May, R. Taniguchi, R. M. Gittelman, M. Klinger, T. M. Snyder, R. Roper, G. Wojciechowska, K. Murray, R. Edmark, S. Evans, L. Jones, Y. Zhou, L. Rowen, R. Liu, W. Chour, H. A. Algren, W. R. Berrington, J. A. Wallick, R. A. Cochran, M. E. Micikas, T. Wrin, C. J. Petropoulos, H. R. Cole, T. D. Fischer, W. Wei, D. S. B. Hoon, N. D. Price, N. Subramanian, J. A. Hill, J. Hadlock, A. T. Magis, A. Ribas, L. L. Lanier, S. D. Boyd, J. A. Bluestone, H. Chu, L. Hood, R. Gottardo, P. D. Greenberg, M. M. Davis, J. D. Goldman and J. R. Heath (2022). “Multiple early factors anticipate post acute COVID-19 sequelae.” Cell 185(5): 881–895.e820.

60. Synolaki, E., V. Papadopoulos, G. Divolis, O. Tsahouridou, E. Gavriilidis, G. Loli, A. Gavriil, C. Tsigalou, N. R. Tziolos, E. Sertaridou, B. Kalra, A. Kumar, P. Rafailidis, A. Pasternack, D. T. Boumpas, G. Germanidis, O. Ritvos, S. Metallidis, P. Skendros and P. Sideras (2021). “The Activin/Follistatin Axis Is Severely Deregulated in COVID-19 and Independently Associated With In-Hospital Mortality.” The Journal of Infectious Diseases 223(9): 1544–1554.

61. Titze-de-Almeida, R., T. R. da Cunha, L. D. Dos Santos Silva, C. S. Ferreira, C. P. Silva, A. P. Ribeiro, A. de Castro Moreira Santos Junior, P. R. de Paula Brandao, A. P. B. Silva, M. C. O. da Rocha, M. E. Xavier, S. S. Titze-de-Almeida, H. E. Shimizu and R. N. Delgado-Rodrigues (2022). “Persistent, new-onset symptoms and mental health complaints in Long COVID in a Brazilian cohort of non-hospitalized patients.” BMC Infect Dis 22(1): 133.

62. Tretter, Y. P., M. Hertel, B. Munz, G. ten Bruggencate, S. Werner and C. Alzheimer (2000). “Induction of activin A is essential for the neuroprotective action of basic fibroblast growth factor in vivo.” Nat Med 6(7): 812–815.

63. Trier, N. H., B. E. Holm, J. Heiden, O. Slot, H. Locht, H. Lindegaard, A. Svendsen, C. T. Nielsen, S. Jacobsen, E. Theander and G. Houen (2018). “Antibodies to a strain-specific citrullinated Epstein-Barr virus peptide diagnoses rheumatoid arthritis.” Scientific Reports 8(1): 3684.

64. Twisk, F. N. and M. Maes (2009). “A review on cognitive behavorial therapy (CBT) and graded exercise therapy (GET) in myalgic encephalomyelitis (ME) / chronic fatigue syndrome (CFS): CBT/GET is not only ineffective and not evidence-based, but also potentially harmful for many patients with ME/CFS.” Neuro Endocrinol Lett 30(3): 284–299.

65. Vojdani, A., E. Vojdani, E. Saidara and M. Maes (2023) “Persistent SARS-CoV-2 Infection, EBV, HHV-6 and Other Factors May Contribute to Inflammation and Autoimmunity in Long COVID.” Viruses 15 DOI: 10.3390/v15020400.

66. Williams, M. V., B. Cox and M. E. Ariza (2016). “Herpesviruses dUTPases: A New Family of Pathogen-Associated Molecular Pattern (PAMP) Proteins with Implications for Human Disease.” Pathogens 6(1).

67. Yang, C., H. Zhao, E. Espín and S. J. Tebbutt (2023). “Association of SARS-CoV-2 infection and persistence with long COVID.” Lancet Respir Med 11(6): 504–506.

68. Zachrisson, O., B. Regland, M. Jahreskog, M. Kron and C. G. Gottfries (2002). “A rating scale for fibromyalgia and chronic fatigue syndrome (the FibroFatigue scale).” Journal of Psychosomatic Research 52(6): 501–509.

69. Zheng, F., A. Link and C. Alzheimer (2017). “Role of activin in cognitive functions, affective behavior and neuronal survival.” 23(2): 85–92.

70. Zubchenko, S., I. Kril, O. Nadizhko, O. Matsyura and V. Chopyak (2022). “Herpesvirus infections and post-COVID-19 manifestations: a pilot observational study.” Rheumatology International 42(9): 1523–1530.

